# Genetically-predicted vitamin D status, ambient UVB during the pandemic and COVID-19 risk in UK Biobank: Mendelian Randomisation study

**DOI:** 10.1101/2020.08.18.20177691

**Authors:** Xue Li, Jos van Geffen, Michiel van Weele, Xiaomeng Zhang, Yazhou He, Xiangrui Meng, Maria Timofeeva, Harry Campbell, Malcolm Dunlop, Lina Zgaga, Evropi Theodoratou

## Abstract

A growing body of evidence shows that poor vitamin D status has been associated with an increased susceptibility to viral and bacterial respiratory infections. In this study, we aimed to examine the association between vitamin D and COVID-19 risk and outcomes, and to explore potential causal effects. We used logistic regression to identify associations between different vitamin D variables (25-hydroxyvitamin D concentration (25-OHD), ambient UVB and genetically-predicted 25-OHD concentrations) and COVID-19 (risk of infection, hospitalisation and death) in 495,780 participants from UK Biobank. We subsequently performed a Mendelian Randomisation (MR) study to test if there was any causal effect. In total, 1,746 COVID-19 cases and 399 COVID-19 deaths occurred between March and June 2020. We found significant inverse associations between COVID-19 infection and 25-OHD in univariable models, but these associations were non-significant after adjustment for confounders. Ambient UVB was strongly and inversely associated with hospitalization and death. Although the main MR analysis showed that genetically-predicted vitamin D levels were not causally associated with COVID-19 risk, MR sensitivity analysis using weighted mode method indicated a potential causal effect (OR=0.72, 95% CI:0.53-0.98; P=0.041). In conclusion, our study found suggestive evidence of association between vitamin D and the risk or severity of COVID-19 but further studies are needed.

## Introduction

Vitamin D is known to improve the function of immune cells, including T-cells and macrophages, and thus plays a critical role in promoting immune responses (1). It has been found to have both antiinflammatory and immune-regulatory properties, which are crucial for the purposeful activation of the immune system defence mechanism (2). A growing body of evidence shows that poor vitamin D status might be associated with an increased susceptibility to viral and bacterial respiratory infections (3, 4).

By analysing publicly available patient data, researchers have discovered a strong correlation between vitamin D deficiency and COVID-19 risk (5). It has also been noted that patients from countries with high COVID-19 mortality rates, such as Italy, Spain and the UK, had lower levels of vitamin D compared to patients in countries that were not as severely affected such as South Korea (5). Furthermore, there is evidence that COVID-19 disproportionately affects black and minority ethnic individuals, with one potential explanation – in addition to other risk factors – being the higher prevalence of vitamin D deficiency (6). It is thus hypothesised that having adequate vitamin D levels may help reduce the risk of contracting the SARS-CoV-2 virus, or reduce the risk of severe or lethal COVID-19 disease.

The main aim of the current study is to perform Mendelian Randomisation (MR) analyses investigating the effect of genetically-predicted vitamin D levels on COVID-19 risk while taking into account ambient UVB radiation at the time of the pandemic, and compare these findings with results obtained from the observational analysis.

## Methods

### Data sources

Basic demographic information and genotype data on 495,780 participants from UK Biobank (7), a large prospective study, were linked to COVID-19 test results (for the period 16/03/2020 to 29/06/2020 provided by Public Health England), including the specimen date, origin (whether the person was an inpatient or not) and result (positive or negative), and death cases caused by clinical and epidemiological diagnosed COVID-19 from death registry. Confirmed COVID-19 cases were defined as UK Biobank participants who had at least one positive test result or died of COVID-19. Participants who have not been tested for SARS-CoV-2 or did not die from COVID-19 were taken as controls. Participants who tested negative were excluded from being controls. Total plasma 25-hydroxy-Vitamin D (25-OHD) was measured at the baseline assessment visits between 2006 and 2010 (median: 11 years before COVID-19), using immunoassay (Diasorin). To remove the effect of sampling season on 25-OHD levels, we generated May-standardised 25-OHD levels (approximating 25-OHD concentration if blood was drawn in May), by applying coefficients generated in a model restricted to controls and adjusted for age and sex (8). Vitamin D status was further categorised as deficient (25-OHD<25 nmol/L), insufficient (25-50 nmol/L), or sufficient (>50 nmol/L). A total of 138 genetic variants have recently been reported to be associated with vitamin D from the largest Genome Wide Association Study (GWAS; n = 443,734) (9). We excluded ambiguous AT and CG variants (n = 4, rs184958517, rs200641845, rs529640451, rs536006581) to avoid bias due to strand differences between studies, and finally 134 SNPs were selected as genetic instruments for the MR analysis. A weighted genetic risk score (wGRS) was calculated as a proxy of genetically-predicted 25-OHD levels for a life-long exposure by using effect estimates reported by Manousaki et al (9).

Dermal synthesis following exposure to UVB radiation is a major source of vitamin D for humans. We used ambient UVB radiation to approximate vitamin D status attributable to dermal synthesis (vitD-UVB) at the time of COVID-19 diagnosis. To do this, we calculated the cumulative and weighted vitD-UVB dose form the TEMIS database, version 2.0 (http://www.temis.nl/uvradiation/UVdose.html), as it has been done previously (10–12). Briefly, we extracted daily UVB dose at wavelengths that induce vitamin D synthesis at each participant’s residential location over 135 days preceding the date of diagnosis for cases. Dates were randomly allocated to controls, from the distribution that was identical to that observed in cases. We weighted the daily UVB contributions before summing them up because more recent UVB exposure contributes more than exposures from a more distant past, since vitamin D is being synthesised and used up. More details on the method can be found elsewhere (10–12).

Additionally, five variants (rs7975232, rs1544410, rs2228570, rs731236 and rs11568820) that are associated with vitamin D receptor (VDR) function were tested for any effect modification on the association between vitamin D and COVID-19 risk, considering VDR mediates biological effects of vitamin D.

### Statistical analysis

In the descriptive analysis, mean and Standard Deviation (SD) is given for continuous variables, and number (N) and proportion for categorical variables, unless indicated otherwise. Logistic regression modelling was used to estimate the effect of vitamin D variables on COVID-19 risk (the risk of infection, hospitalisation and death), in unadjusted models and after adjustment for a range of confounders, including age, sex, body mass index (BMI), month of blood draw, ethnicity (i.e., White, Asian, Black and others), physical activity (number of days for moderate/vigorous/walked activity), smoking and alcohol status (i.e., current, previous, never, unknown), sunshine exposure variables (i.e., time spend outdoors in summer, time spent outdoors in winter and the use of sun/uv protection) and vitamin D supplement intake. Specifically, we investigated the associations between COVID-19 and: (i) vitamin D levels (circulating 25-OHD concentration, May-standardised 25-OHD concentration, and categorical vitamin D status); (ii) vitD-UVB predicted by an integrative measure of ambient UVB radiation at the time of the pandemic; (iii) genetically-predicted 25-OHD concentration using wGRS (vitD-wGRS_134_), in an unadjusted model and fully adjusted models as described above. We also tested their interactions with VDR SNPs. For MR analyses, vitamin D SNPs were aligned by their effect alleles; inverse-variance weighted MR approach (13) was used as the main analysis, and the simple mode, Egger, weighted median and weighted mode as sensitivity analyses (14) to explore the robustness of the findings in the presence of potential pleiotropy of the genetic variants. All analyses were conducted using R version 3.6.1.

## Results

There was a total of 14,439 COVID-19 tests conducted for 7,898 UK Biobank participants. Of these, 1,596 individuals had at least one positive COVID-19 test and 1,020 of them were hospitalised. Additional 399 COVID-19 death cases were identified from the death registry. **Table 1** presents the basic demographic characteristics of the cohort. The mean age of the COVID-19 cases (including deaths) was 68.8y ± 9.2y, and 52.9% were male. Approximately, 86.5% cases were white, 4.8% Asian, 2.8% black, and 5.8% other. The proportion of vitamin D supplement intake in controls (4.2%) was relatively higher than that in COVID-19 cases (4.0%). Median 25-OHD concentration measured at recruitment was slightly lower in COVID-19 cases (46.57 (IQR 29.70-60.83) nmol/L) than controls (46.90 (IQR 32.40-62.40) nmol/L), more so among those who died (44.3 (IQR 29.7-60.42) nmol/L). The ambient UVB relevant for vitamin D synthesis (vitD-UVB) was also lower in cases 65.95 kJ/m^2^ (IQR 38.10-99.08 kJ/m^2^) than in controls 66.14 kJ/m^2^ (IQR 37.61-100.50 kJ/m^2^), and it was much lower for death cases (43.09 kJ/m^2^ (IQR 31.89-74.10 kJ/m^2^)). In univariate analysis, vitD-UVB at recruitment was strongly associated with 25OHD concentrations at recruitment (beta = 0.10, p-val < 2 ×10^−16^, R^2^ = 0.12) and also in multivariate model (beta = 0.11, p-val < 2 ×10^−16^, R^2^ = 0.19). The variance of 25OHD concentration at recruitment explained by vitD-UVB at recruitment alone was 12.4%, by vitD-GRS_134_ alone was 4.2%, and by vitD-GRS_134_ and vitD-UVB together with covariates in a multivariate model was 23.1%.

**Table 1:** Baseline characteristics of the COVID-19 cases and controls in UK Biobank.

|  | COVID-19 cases |  |  | Total cases (n=1746) | Controls (n=494,034) |
| --- | --- | --- | --- | --- | --- |
|  | Outpatient (n=576) | Inpatient (n=1020) | Death (n=399) |  |  |
| Gender, N (%) | Male: 264 (45.8%)<br>Female: 312 (54.2%) | Male: 570 (55.9%)<br>Female: 450 (44.1%) | Male: 255 (63.9%)<br>Female: 144 (36.1%) | Male: 924 (52.9%)<br>Female: 822 (47.1%) | Male: 268,549 (54.5%)<br>Female: 224,485 (45.5%) |
| Age, Mean (SD) | 65.97 (9.42) | 69.40 (8.86) | 74.66 (5.98) | 68.76 (9.18) | 68.44 (8.11) |
| BMI (kg/m<sup>2</sup>) | 27.66 (5.01) | 27.47 (4.81) | 27.59 (4.68) | 27.53 (4.88) | 27.42 (4.79) |
| vitD (nmol/L), median (IQR) | 46.11 (30.20-59.30) | 46.82 (29.40-61.35) | 44.30 (29.70-60.42) | 46.57 (29.70-60.83) | 46.90 (32.40-62.40) |
| vitD-UVB (kJ/m<sup>2</sup>), median (IQR) | 90.80 (67.43-98.95) | 48.96 (33.81-81.08) | 43.09 (31.89-74.10) | 65.95 (38.10-99.08) | 66.14 (37.61-100.50) |
| Vitamin D supplement, N (%) | 21 (3.6%) | 43 (4.2%) | 12 (3.0%) | 64 (4.0%) | 20,861 (4.2%) |
| Smoking status, N (%) |  |  |  |  |  |
| Never | 318 (55.2%) | 462 (45.3%) | 148 (37.1%) | 780 (48.9%) | 269,014 (54.5%) |
| Previous | 190 (33.0%) | 429 (42.0%) | 186 (46.6%) | 619 (38.8%) | 169,392 (34.3%) |
| Current | 63 (10.9%) | 116 (11.4%) | 60 (15.0%) | 179 (11.2%) | 51,748 (10.5%) |
| Unknown | 5 (0.9%) | 13 (1.3%) | 5 (1.3%) | 18 (1.1%) | 3,880 (0.8%) |
| Alcohol drinker status, N (%) |  |  |  |  |  |
| Never | 45 (7.8%) | 79 (7.7%) | 30 (7.5%) | 124 (7.8%) | 21,988 (4.4%) |
| Previous | 22 (3.8%) | 61 (6.0%) | 31 (7.8%) | 83 (5.2%) | 17,637 (3.6%) |
| Current | 508 (88.2%) | 873 (85.6%) | 334 (83.7%) | 1381 (86.5%) | 451,954 (91.5%) |
| Unknown | 1 (0.2%) | 7 (0.7%) | 4 (1.0%) | 8 (0.5%) | 2,455 (0.5%) |
| Ethnicity, N (%) |  |  |  |  |  |
| White | 497 (86.3%) | 883 (86.6%) | 357 (89.5%) | 1380 (86.5%) | 464,012 (93.9%) |
| Asian | 21 (3.6%) | 56 (5.5%) | 11 (2.8%) | 77 (4.8%) | 11,150 (2.3%) |
| Black | 17 (3.0%) | 28 (2.7%) | 12 (3.0%) | 45 (2.8%) | 4,492 (0.9%) |
| Other/Unknown | 41 (7.1%) | 53 (5.2%) | 19 (4.7%) | 94 (5.9%) | 14,380 (2.9%) |
| Month (of COVID-19 diagnosis) |  |  |  |  |  |
| March | 47 (8.2%) | 257 (25.2%) | 114 (28.6%) | 328 (18.8%) | 93184 (18.9%) * |
| April | 242 (42.0%) | 563 (55.2%) | 228 (57.1%) | 882 (50.5%) | 248489 (50.4%) * |
| May | 218 (37.8%) | 161 (15.8%) | 49 (12.3%) | 420 (24.1%) | 118328 (24.0%) * |
| June | 69 (12.0%) | 39 (3.8%) | 8 (2.0%) | 116 (6.6%) | 33033 (6.7%) * |

\*Dates were randomly allocated to controls for the calculation of vitD-UVB based on the distribution that was identical to that observed in cases.

Results from logistic regression analysis **(Table 2)** showed significant associations between COVID-19 infection and vitamin D levels (both crude and May-standardised 25OHD concentration) in univariable models (OR = 0.995, 95%CI:0.992-0.997, p-val = 2.05×10^”5^ and OR = 0.993, 95%CI:0.991-0.996, p-val = 2.05 ×10^”6^, respectively); however, these associations were not significant after adjustment for covariates (OR = 1.00, 95%CI:0.99-1.01, P = 0.573 and OR = 1.00, 95%CI:0.99-1.01, P = 0.578, respectively). Compared to vitamin D deficient individuals, the risk of infection was 16% and 31% lower in those who were insufficient or sufficient, respectively, in univariable models, but not in multivariable models. Neither vitD-UVB nor genetically-predicted (vitD-GRS_134_) vitamin D were associated with COVID-19 infection risk. In order to investigate whether vitamin D levels would influence COVID-19 severity, we performed a sensitivity analysis for hospitalised cases and COVID-19 deaths. We consistently found that vitD-UVB dose was strongly and inversely associated with the hospitalization and death from COVID-19 in univariable and multivariable models (OR = 0.98, 95%CI: 0.97-0.99, p-val < 2×10^−16^), while null findings were reported for other vitamin D variables **(Table 2)**.

**Table 2:** Association between Vitamin D and COVID-19 risk.

|  | COVID positive (N=1746) vs controls (N=494,034) |  |  |  | COVID hospitalization (N=1020) vs non- hospitalization (N=576) cases |  |  |  | COVID death (N=399) vs non-death (N=1347) cases |  |  |  |
| --- | --- | --- | --- | --- | --- | --- | --- | --- | --- | --- | --- | --- |
|  | Univariable |  | Multivariable<sup>a</sup> |  | Univariable |  | Multivariable<sup>a</sup> |  | Univariable |  | Multivariable<sup>a</sup> |  |
|  | OR (95% CI) | p-val | OR (95% CI) | p-val | OR (95% CI) | p-val | OR (95% CI) | p-val | OR (95% CI) | p-val | OR (95% CI) | p-val |
| vitD (nmol/L)<sup>†</sup> | 0.995 (0.992-0.997) | 2.05×10<sup>-5</sup> | 1.00 (0.99-1.01) | 0.573 | 1.00 (0.99-1.01) | 0.556 | 1.00 (0.99-1.01) | 0.608 | 1.00 (0.99-1.01) | 0.994 | 1.00 (0.99-1.01) | 0.687 |
| vitD_May_adjusted (nmol/L)<sup>†</sup> | 0.993 (0.991-0.996) | 2.09×10<sup>-6</sup> | 1.00 (0.99-1.01) | 0.578 | 1.00 (0.99-1.01) | 0.615 | 1.00 (0.99-1.01) | 0.784 | 1.00 (0.99-1.01) | 0.360 | 1.00 (0.99-1.01) | 0.770 |
| vitD_categorical<sup>†</sup> |  |  |  |  |  |  |  |  |  |  |  |  |
| 0-25 nmol/L | ref | -- | ref | -- | ref | -- | ref | -- | ref | -- | ref | -- |
| 25-50 nmol/L | 0.84 (0.73-0.97) | 0.020 | 0.97 (0.83-1.13) | 0.658 | 0.83 (0.61-1.14) | 0.262 | 0.88 (0.62-1.25) | 0.483 | 0.82 (0.58-1.16) | 0.269 | 0.78 (0.52-1.16) | 0.222 |
| 50 nmol/L | 0.69 (0.60-0.81) | 1.22×10<sup>-6</sup> | 0.88 (0.74-1.05) | 0.153 | 0.93 (0.68-1.29) | 0.676 | 0.91 (0.62-1.34) | 0.634 | 0.94 (0.66-1.33) | 0.717 | 0.83 (0.54-1.29) | 0.406 |
| vitD-wGRS<sub>134</sub> | 0.90 (0.79-1.02) | 0.105 | 0.92 (0.81-1.04) | 0.197 | 1.05 (0.82-1.34) | 0.705 | 1.04 (0.81-1.35) | 0.740 | 1.33 (0.98-1.78) | 0.062 | 1.27 (0.93-1.74) | 0.137 |
| vitD-UVB<sup>§</sup> | 1.00 (0.99-1.01) | 0.444 | 1.00 (0.99-1.01) | 0.415 | 0.98 (0.97-0.99) | <2×10<sup>-16</sup> | 0.98 (0.97-0.99) | <2×10<sup>-16</sup> | 0.98 (0.97-0.99) | <2×10<sup>-16</sup> | 0.98 (0.97-0.99) | <2×10<sup>-16</sup> |
| **vitD-wGRS<sub>134</sub> + vitD-UVB** |  |  |  |  |  |  |  |  |  |  |  |  |
| vitD-wGRS<sub>134</sub> | 0.90 (0.80-1.03) | 0.124 | 0.93 (0.82-1.06) | 0.282 | 0.87 (0.66-1.14) | 0.306 | 0.86 (0.65-1.14) | 0.298 | 1.21 (0.88-1.64) | 0.239 | 1.12 (0.81-1.57) | 0.498 |
| vitD-UVB | 1.00 (0.99-1.01) | 0.461 | 1.00 (0.99-1.01) | 0.373 | 0.98 (0.97-0.99) | <2×10<sup>-16</sup> | 0.98 (0.97-0.99) | <2×10<sup>-16</sup> | 0.98 (0.97-0.99) | <2×10<sup>-16</sup> | 0.98 (0.97-0.99) | <2×10<sup>-16</sup> |

<sup>a</sup> Adjusted for age, gender, Body Mass Index (BMI), month of blood draw (adjusted for vitD and vitD-categorical only), ethnicity, physical activity, smoking and alcohol status, sunshine exposure variables (i.e., time spend outdoors in summer, time spent outdoors in winter and the use of sun/uv protection) and vitamin D supplement intake.

Similar results were observed when we stratified the cohort by BMI (< 25 or ≥ 25) **(Table S1)**. Moreover, in overweight/obese individuals both vitD-UVB and genetically-predicted levels were inversely associated with hospitalisation. When stratifying the study sample by ethnicity, we found that lower vitD-UVB was associated with increased risk of hospitalization and death of COVID-19 cases among White population in both univariable and multivariable models **(Table S2)**. Additionally, in White population, the risk of hospitalisation was around 40% lower in vitamin D sufficient individuals than those who were deficient, in both univariable (p-val = 0.012) and multivariable (p-val = 0.024) models. Although numbers were small, vitD-UVB was also significantly associated with hospitalisations in Asians and with COVID-19 risk in Black population **(Table S3-S4)**. We observed no evidence of interaction between vitD-GRS_134_ and VDR SNPs, or between vitD-UVB and VDR SNPs **(Table S5)**, while evidence of significant interaction was observed between vitD-GRS_134_ and vitD-UVB (beta = –0.005, p-val = 0.001) for COVID-19 infection risk **(Table S6)**. When stratified by vitD-UVB tertiles, the beta-coefficient for vitD-wGRS_134_ changed from 0.179 (p-val = 0.16) in Tertile 1, to −0.161 (p-val = 0.158) in Tertile 2, and was statistically significant in Tertile 3 (beta = –0.321, p-val = 0.004, **Table S7**). **Table S8** shows the results of the MR analyses using 134 genetic instruments, which are also graphically presented in **Figure 1**. The main IVW MR showed that genetically-predicted vitamin D levels were not causally associated with COVID-19 infection **(Table S8)** and null results were obtained in MR sensitivity analyses except for the weighted mode MR, which suggested a potential causal effect (OR = 0.72, 95CI:0.53-0.98; p-val = 0.041).

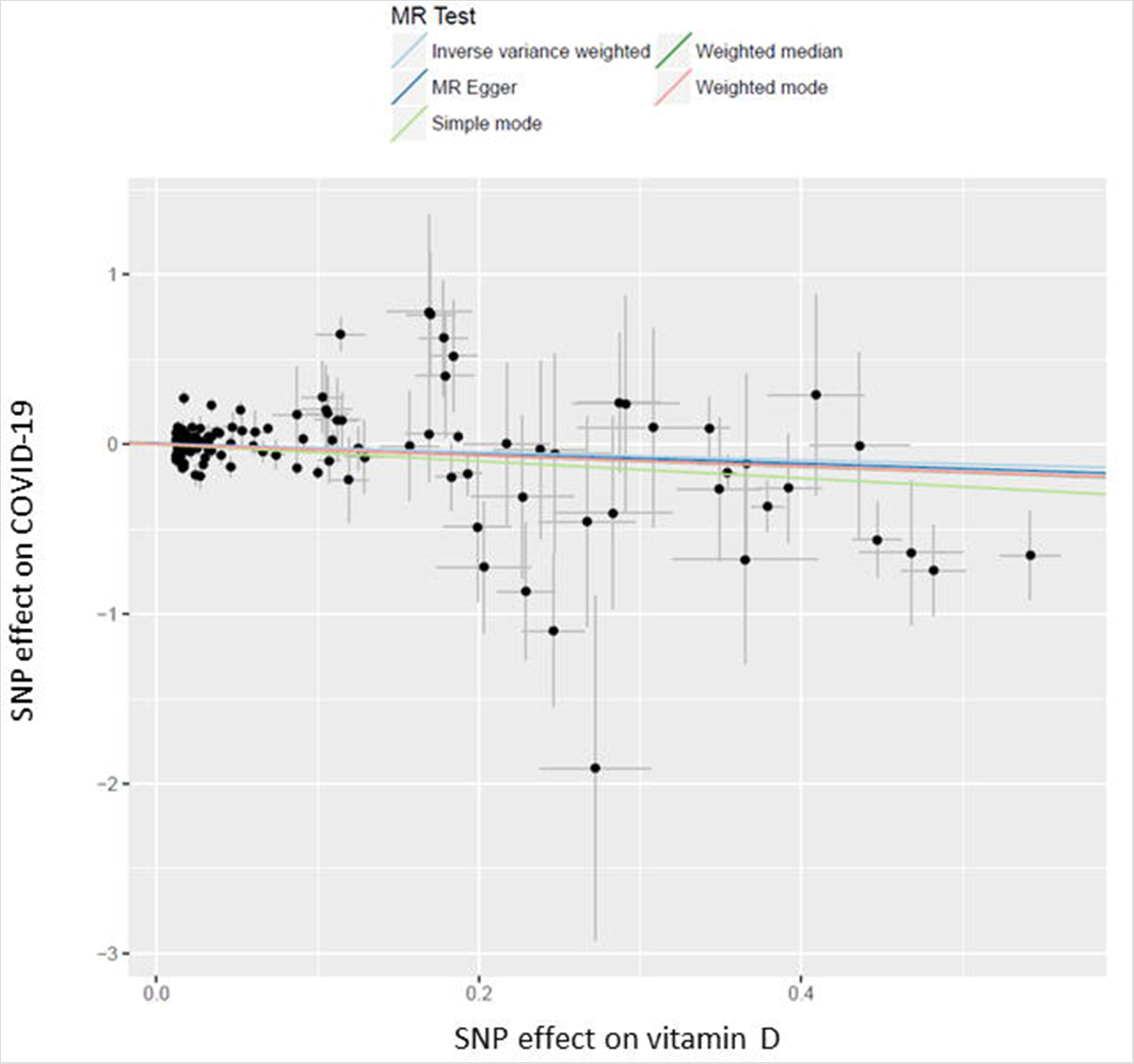

## Discussion

In this study, we assessed whether there is an association between vitamin D and COVID-19 risk and severity by examining a comprehensive set of key vitamin D variables jointly for the first time, and applying a number of analyses to probe consistency of our findings. We found no strong association between vitamin D levels (plasma concentration measured at recruitment, 11 years ago) and COVID-19 risk or severity after adjustment for confounders, results that are in accordance to the recent study by Hastie et al (15). However, we consistently found a strong inverse association between an integrated measure of ambient UVB preceding disease onset (vitD-UVB) and hospitalisation and death among cases. In this cohort, vitD-UVB explained the largest portion of the variance in 25-OHD: vitD-UVB alone explained 12.4%, while vitD-GRS_134_ alone explained 4.2%. Previous studies have shown that heritability of 25-OHD is high in winter and low in summer, which suggests a varied role of genetic factors, dependant on the UVB intensity (16). It is therefore not surprising that we found evidence of an interaction between vitD-UVB and vitamin D genetic risk score, and these findings highlight the added value of examining genetically-predicted levels and ambient UVB jointly. MR sensitivity analysis using the weighted mode method indicated a potential causal effect, although the main MR analysis showed that genetically-predicted vitamin D levels were not causally associated with COVID-19 risk. As COVID-19 test results (inpatient or outpatient) were only available from England, we did a sensitivity analysis after excluding participant from Scotland or Wales and individuals who died before Jan 2020 (since they could not have been infected by SARS-COV-2) from the controls and the results remained unchanged.

Our study has a number of strengths and limitations. Firstly, UK Biobank is a large prospective study, with rich information on a range of demographic, lifestyle and health-related risk factors. Vitamin D plasma measurements were conducted in a single central processing laboratory using the Diasorin immunoassay, albeit a blood sample was taken over a decade ago and is unlikely to be representative of participants’ vitamin D status at the time of the pandemic. We have partially addressed this by using genetic instruments (that are determined by DNA sequence and hence not variable) to derive genetically-predicted vitD levels. It is important to note that heritability of vitamin D status is high in winter (70-90%), but levels might be entirely determined by environmental factors in the summer (16). Therefore, we also included an integrative measure of ambient UVB radiation during the pandemic. However, the discriminatory power of the UVB variable is somewhat limited in this study, because UVB radiation is low at this time of the year, particularly at the high northern latitude of UK. We only used ambient UVB, and did not capture individual behavioural differences that would determine the actual level of vitamin D synthesis in the skin, such as time and time of day spent outside, clothing, choosing to walk on the sunny side of the street;. Moreover, time of year is the strongest predictor of vitD-UVB but control dates were assigned to follow the same distribution as case dates, which consequently might have led to artificially diminished differences in vitD-UVB between cases and controls, but analysis relating to hospitalisation and death are not affected by this. We also conducted an analysis of the genetically-predicted vitamin D and a number of state-of-the-art MR analyses. However, the main limitation is the lack of power (only 46% power for an effect estimate (OR) of 1.2 at a p-value level of 0.05) given the small number of COVID-19 patients and the relatively small percentage of variance (4.2%) explained by vitamin D-related genetic variants. It should be noted that the number of COVID-19 cases in each model were slightly different due to missing variables for vitamin D and genotype data in some cases. Another limitation relates to the fact that not all participants have been tested for present (or past) Covid-19 infection; consequentially, a number of controls may have in fact been cases, further driving our findings to the null. It should be acknowledged that the COVID-19 cases in UK biobank have a high rate of hospitalisation due to the very limited and targeted testing at this stage of the pandemic in the UK, so this study reflects mainly those with more severe COVID-19 and gives less information about true infection risk, or risk of milder disease.

In conclusion, we found some suggestive evidence of an association between vitamin D and risk or severity of COVID-19, which may have implications if a second wave occurs during the autumn or winter months. Further studies are needed to examine the role of vitamin D in COVID-19.

## Data Availability

Details of genotyping and quality control of UK biobank available at: http://biobank.ndph.ox.ac.uk/crystal/docs/genotyping_qc.pdf

Details of genotype imputation of UK biobank available at: http://www.ukbiobank.ac.uk/wpcontent/uploads/2014/04/imputation%20documentation%20May2015.pdf

Participants description of UK biobank available at: https://www.biorxiv.org/content/biorxiv/early/2017/07/20/166298.full.pdf

## Abbreviations

25-OHD: 25-hydroxyvitamin D;
BMI: body mass index;
COVID-19: Corona Virus Disease 2019;
IQR: interquartile range;
MR: Mendelian Randomisation;
OR: Odds Ratio;
SD: Standard Deviation;
UVB: ultraviolet radiation b;
vitD: vitamin D;
wGRS: weighted genetic risk score.

## Acknowledgements

The authors would like to thank all participants in the UK Biobank.

## Availability of data and materials

Details of genotyping and quality control of UK biobank available at: http://biobank.ndph.ox.ac.uk/crystal/docs/genotvping_qc.pdf

Details of genotype imputation of UK biobank available at: http://www.ukbiobank.ac.uk/wpcontent/uploads/2014/04/imputationdocumentationMay2015.pdf

## Authors’ contributions

E.T. and L.Z. conceived the study. X.L. performed the data analysis with input from X.Z., Y.H., X.M. and M.T.; J.G. and M.W. contributed to the calculation of cumulative and weighted vitD-UVB dose. X.L. drafted the manuscript with input from E.T., L.Z.; H.C. and M.D. All authors critically reviewed the manuscript and contributed important intellectual content. All authors have read and approved the final manuscript as submitted.

## Ethics approval and consent to participate

UK Biobank has approval from the North West Multi-Centre Research Ethics Committee (11/NW/0382) and obtained written informed consent from all participants prior to the study. Data used in this study were obtained from UK Biobank under an approved data request application (application ID: 10775). No consent for publication was required.

